# Antipsychotic Polypharmacy and Adverse Drug Reactions Among Adults in a London Mental Health Service, 2008-2018

**DOI:** 10.1101/2021.11.28.21266870

**Authors:** Justin C. Yang, Johan H. Thygesen, Nomi Werbeloff, Joseph F. Hayes, David P.J. Osborn

## Abstract

**Background:** Antipsychotic polypharmacy (APP) occurs commonly but it is unclear whether it is associated with an increased risk of adverse drug reactions. Electronic health records (EHRs) offer an opportunity to examine APP using real-world data. In this study, we use EHR data to identify periods when patients were prescribed 2+ antipsychotics and compare these with periods of antipsychotic monotherapy. To determine the relationship between APP and subsequent instances of adverse drug reactions: QT interval prolongation, hyperprolactinaemia, and increased body weight (body mass index [BMI] ≥ 25).

**Methods:** We extracted anonymised EHR data. Patients aged 16+ receiving antipsychotic medication at Camden & Islington NHS Foundation Trust between 1 January 2008 and 31 December 2018 were included. Multilevel mixed-effects logistic regression models were used to elucidate the relationship between APP and the subsequent presence of QT interval prolongation, hyperprolactinaemia, and/or increased BMI following a period of APP within 7, 30, or 180 days respectively.

**Results:** We identified 35,409 observations of antipsychotic prescribing among 13,391 patients. APP was associated with a subsequent increased risk of hyperprolactinaemia (adjusted odds ratio 2.46; 95% C.I. 1.87-3.24) and of having a BMI > 25 (adjusted odds ratio 1.75; 95% C.I. 1.33-2.31) in the period following the APP prescribing.

**Conclusions:** Our observations suggest that APP should be carefully managed with attention to hyperprolactinaemia and obesity.

## Introduction

The potential adverse effects of antipsychotic use are well-recognised and include QT interval prolongation and torsade de pointes (a specific type of abnormal heart rhythm) (Glassman and Bigger Jr, 2001), hyperprolactinaemia (Peuskens *et al*., 2014), and weight gain (Correll *et al*., 2011). Yet, by comparison, antipsychotic polypharmacy (APP), the common practice of co-prescribing two or more antipsychotics, and the potential adverse effects arising from this practice are understudied. Previous research has not found an increased risk of mortality with APP as compared to antipsychotic monotherapy, though research has been largely heterogeneous with variations in both methodological approaches and definitions of APP (Kadra *et al*., 2018b; Taipale *et al*., 2018; Kasteridis *et al*., 2019; Buhagiar *et al*., 2020). The relationship between APP and QT interval appears to be inconclusive, with a meta-analysis by Takeuchi et al. 2015, pointing out that the evidence is scarce and inconsistent (Takeuchi *et al*., 2015). Some evidence suggests that APP might be linked to hyperprolactinaemia and weight gain, but again the evidence is inconclusive as some studies also find weight loss when comparing patients on APP with monotherapy (Gallego *et al*., 2012). While in certain cases there may be a clear clinical indication and benefit in the co-prescription of specific antipsychotic medications (e.g. better symptom control with clozapine plus another antipsychotic), and mitigating metabolic side-effects with a concomitant use of aripiprazole (Fleischhacker and Uchida, 2014), the general practice of APP should be the exception rather than the rule, due to the limited knowledge of potential adverse drug reactions (ADRs), and the general finding that the overall global burden of ADRs increases with APP (Gallego *et al*., 2012). The heterogeneity in study designs of previous studies, variations in definitions of APP, and conflicting results among prior work highlights the need for a stronger evidence base in this area, especially drawing upon real-world evidence to better inform clinical practice. Randomized controlled trials (RCTs), in particular, have often only examined the practice of APP with relatively brief follow-up, suggesting a need for further research (Buhagiar *et al*., 2020). Increasingly, routinely collected electronic health record (EHR) data have been leveraged to conduct large, high-quality studies which can address research questions which are otherwise difficult to approach using RCTs or conventional cohort designs. These studies can draw upon the wealth of real world clinical data recorded from interactions with patients, using both structured fields, which use well-defined taxonomies, as well as clinical free-text via natural language processing (NLP). In psychiatry, more specifically, these approaches have previously been used to investigate the efficacy of individual antipsychotics (Patel *et al*., 2018), rates of long-term APP among patients with severe mental illness (Kadra *et al*., 2015), and risk of readmission following APP (Kadra *et al*., 2018a).

### Aims of the Study

This study aims to use routinely collected EHR data to identify instances of antipsychotic polypharmacy (APP) in a large NHS mental health provider and to investigate association between APP and the subsequent presence of three specific adverse drug reactions (ADRs): prolonged corrected QT interval (QTc), hyperprolactinaemia, and increased body mass index (BMI). We hypothesised that ADRs would be associated with periods on or following APP due to the potentially additive harmful effects of taking multiple antipsychotics at the same time.

## Method

### Participants

Camden & Islington NHS Foundation Trust (C&I NHS FT) provides secondary mental health services to around 470,000 individuals in the London boroughs of Camden and Islington (Werbeloff *et al*., 2018). We examined patients aged 16 and above who had contact with C&I NHS FT between 1 January 2008 and 31 December 2018. During this period, the C&I NHS FT Clinical Record Interactive Search (CRIS) research database included anonymised information for approximately 149,000 unique patients receiving care at the Trust. We included patients who were prescribed at least one antipsychotic medication and had at least one measurement of any of the clinical outcomes of interest, namely, QTc, prolactin level, or BMI (Supplementary Table 1).

We extracted data using CRIS (Fernandes *et al*., 2013), an application which allows for anonymisation of routinely collected EHR data, including both structured fields (i.e., gender and ethnicity), and unstructured fields (i.e., clinical documentation and progress notes). Research approval for the C&I NHS FT CRIS research database was obtained from the East of England - Cambridge Central Research Ethics Committee (19/EE/0210), and the project protocol was reviewed and approved by the C&I NHS FT Research Oversight Committee. Individuals who have opted out from being included in CRIS, including those who have opted out through the national data opt-out, were excluded.

### Variables

#### Antipsychotic Prescribing and Polypharmacy

We identified instances of antipsychotic prescribing from clinical free text (including progress notes and letters) through the use of an NLP application for ‘medication’ developed by the South London and Maudsley NHS Foundation Trust (SLaM NHS FT) Biomedical Research Centre (BRC) (Perera *et al*., 2016). The application used a gazetteer of generic and commercial names for all medications in use in the UK to ascertain instances where patients were reported as receiving these, with supplementary rules for ascertaining recorded dose, frequency/timing and starting/stopping statements (CRIS Natural Language Processing, n.d.). As temporal identification was important for our analysis, we only considered recorded instances of prescribed antipsychotics in the present tense. Date stamps from the source documents were used as a proxy for the prescription date. Following extraction, all antipsychotic medications were identified and multiple names for the same compound grouped together and standardised for subsequent analyses (Supplementary Table 2). The reported precision and recall for the NLP medications identification algorithm range from 76-88% and from 69-88%, respectively, based on data from a comparable trust using EHR data at SLaM NHS FT (CRIS Natural Language Processing, n.d.). We validated the results of the NLP medications app on C&I NHS FT data by manually reviewing 100 identified matches on a set of patients, yielding a positive predictive value (PPV) of 98% and a negative predictive value (NPV) of 100%. Moreover, we observed specificity and sensitivity of 100%..

The period during which an antipsychotic was considered prescribed was defined as the contiguous period between the first date on which antipsychotic prescription was identified and the last date on which antipsychotic prescription was identified within the study period. It is possible that an antipsychotic may have continued to be prescribed to a patient following this final observation but this information was not recorded in the EHR, and thus could not be inferred for our analysis.

To distinguish true periods of APP from instances of tapering or transitioning from one antipsychotic to another, APP was defined as the co-prescription of at least two different antipsychotics for a minimum of 30 days. This threshold was determined a priori in consultation with clinicians at Camden & Islington NHS Foundation Trust who conveyed that 30 days would be a clinically meaningful ongoing co-prescription of two or more antipsychotics which would largely exclude periods of co-prescription for brief periods of agitation (e.g. pro re nata [PRN] prescribing) or during cross-titration between two different antipsychotics.

#### Clinical Measurements and Outcomes

Clinical measurements for each of QTc, prolactin and BMI were obtained for all patients aged 16 and over who were prescribed at least one antipsychotic medication between 1 January 2008 and 31 December 2018. Clinical measurements of QTc, prolactin, and BMI were identified from free-text in the EHR by developing regular expressions for the capture of each of the three measures. We used an iterative process to develop the expressions by reviewing their capture and refining the expressions to gain a PPV higher than 95%. We tested the PPV of each regular expression by manually reviewing 100 identified matches on a new unseen set of patients and found PPVs and NPVs of 100% for each of QTc, prolactin and BMI. Furthermore, our regular expressions yielded sensitivities and specificities of 100%. Regular expressions and illustrative examples of matches are shown in Supplementary Table 2.

Prolonged QTc was defined as values above 450ms and 470ms for men and women respectively (Xiong *et al*., 2020). Hyperprolactinemia was defined as prolactin measurements greater than 450 mIU/L for men or 500 mIU/L for women (Peveler *et al*., 2008). Overweight was defined as BMI greater than or equal to 25 (Tsigos *et al*., 2008). Supplementary Figure 1 shows a schematic of how clinical measurements were classified in this study.

#### Exposure Period

Clinically, we expect the time leading to the onset of an ADR following initiation of APP to vary depending on the specific aetiology and biological mechanism of the ADR. For instance, we expect QTc prolongation to be observable shortly after the start of APP, likely within one week (Zareba and Lin, 2003), while changes to BMI may not be observable until months after start of APP (Zhang *et al*., 2016). Consequently, we focused on detected instances of QT interval prolongation, hyperprolactinaemia, and/or increased body weight during or following a period of APP within 7, 30, or 180 days respectively. Supplementary Figure 2 illustrates how clinical measurements relate to periods of polypharmacy and exposure periods in this study. As such, individual patients could contribute clinical measurements both within and outside exposure periods and could contribute multiple measurements during exposed and unexposed periods.

#### Other Covariates

Gender and ethnicity were extracted from structured fields. We derived the patients’ Index of Multiple Deprivation (IMD) for their local area. IMD combines national census information from 38 indicators into seven domains of deprivation, yielding a single deprivation score for 32,482 lower super output areas (LSOAs) in England, each containing approximately 1,500 people or 400 households. Patient’s postcodes are routinely collected as part of care and this information is used to determine a patient’s LSOA, prior to anonymisation. We linked IMD scores by cross-referencing each patient’s LSOA to 2015 IMD national data. These were then separated by tertiles with the lowest category signifying most deprived. Individuals with missing data for these covariates were excluded from this study.

Individual patients who are seen often or are identified as high risk for adverse drug outcomes by clinicians may receive more testing or closer clinical observation than those who do not present with such risk. This may increase the likelihood of identification of ADRs in certain patient groups and risk biasing the independence of the ADR detection across the sample. To account for this, the frequency of service usage was estimated as the number of recorded clinical measurements for the study outcomes. These proxies for service usage were included in the regression analyses examining the associations with APP. We also created a variable for the length of time known to C&I NHS FT in years, but found this variable highly correlated with service usage, and thus to avoid collinearity did not use this variable in our analysis.

### Analysis

Analyses were conducted using R version 4.0.2. Univariable and multivariable multilevel mixed-effects logistic regression models were used to elucidate the relationship between periods of APP and subsequent cases of adverse drug reactions, accounting for clustering of measurements within individual patients (Quené and van den Bergh, 2004).

Univariable and multivariable multilevel mixed-effects logistic regressions were conducted for each of the three ADRs (modelling individual patients as random effects, allowing for the inclusion of individuals at multiple timepoints, both as receiving APP treatment and not, as well as having above and below threshold measures of ADR (see supplementary figure 2). For each ADR, a dichotomous variable indicated whether or not a clinical measurement represented an ADR. The primary predictor variable was whether or not the ADR was recorded during an exposure period. Multivariable regressions also included covariates to adjust for age, gender, total number of clinical measurements, ethnicity, and IMD.

## Results

Table 1 describes the main characteristics of our study sample. We identified 35,409 observations of antipsychotic prescribing among 13,391 patients during the study period (1 January 2008 and 31 December 2018). This represents a total of 9.0% of the full patient population at C&I NHS FT were being prescribed antipsychotic medication at some point in that period. 3,313 (24.7%) of patients prescribed antipsychotics had at least one period of APP lasting at least 30 days. The positive predictive value of the medications NLP app used to detect antipsychotic medications was 98%. Supplementary Figure 3 shows the distribution of observations per individual and Supplementary Figure 4 shows the distribution of duration of polypharmacy (in days).

**Table 1.** Sample demographics. Characteristics for patients who received antipsychotic medication and had at least one measure of corrected QT interval(QTc), prolactin level, or body mass index (BMI) between 1 January 2008 and 31 December 2018. Numbers are stratified by measurements for QTc, prolactin level measurements, or BMI within 7, 30 or 180 days of antipsychotic polypharmacy (APP). Numbers are reported as counts (with percentage) or medians (with interquartile ranges [IQRs]).

|  | ≥1 QTc Measurement<br>During or<br>Within 7 Days of APP |  | ≥1 Prolactin Measurement<br>During or<br>Within 30 Days of APP |  | ≥1 BMI Measurement<br>During or<br>Within 180 Days of APP |  |
| --- | --- | --- | --- | --- | --- | --- |
|  | No | Yes | No | Yes | No | Yes |
| **Number of Patients** | 2313 | 1000 | 862 | 619 | 2748 | 1115 |
| **Median Number of Measurements ± IQR** | 2.0 ± 1.5 | 4.0 ± 3.5 | 1.0 ± 0.5 | 2.0 ± 1.0 | 1.0 ± 1.0 | 2.0 ± 1.5 |
| **Median Age ± IQR** | 47.0 ± 14.5 | 44.0 ± 10.5 | 44.0 ± 12.5 | 43.0 ± 10.5 | 47.0 ± 12.0 | 44.0 ± 10.0 |
| **Gender** |  |  |  |  |  |  |
| **Male** | 1259 (54.4%) | 540 (54.0%) | 449 (52.1%) | 316 (51.1%) | 1399 (50.9%) | 605 (54.3%) |
| **Female** | 1054 (45.6%) | 460 (46.0%) | 413 (47.9%) | 303 (48.9%) | 1349 (49.1%) | 510 (45.7%) |
| **Ethnicity** |  |  |  |  |  |  |
| **White British** | 891 (38.5%) | 365 (36.5%) | 323 (37.5%) | 205 (33.1%) | 1123 (40.9%) | 377 (33.8%) |
| **White Other** | 525 (22.7%) | 177 (17.7%) | 215 (24.9%) | 107 (17.3%) | 597 (21.7%) | 219 (19.6%) |
| **Asian** | 154 (6.7%) | 78 (7.8%) | 51 (5.9%) | 48 (7.8%) | 176 (6.4%) | 98 (8.8%) |
| **Black** | 491 (21.2%) | 293 (29.3%) | 176 (20.4%) | 194 (31.3%) | 534 (19.4%) | 323 (29.0%) |
| **Mixed** | 98 (4.2%) | 42 (4.2%) | 41 (4.8%) | 37 (6.0%) | 120 (4.4%) | 48 (4.3%) |
| **Other** | 154 (6.7%) | 45 (4.5%) | 56 (6.5%) | 28 (4.5%) | 198 (7.2%) | 50 (4.5%) |
| **IMD Tertile** |  |  |  |  |  |  |
| **1 (Most Deprived)** | 1633 (70.6%) | 725 (72.5%) | 574 (66.6%) | 425 (68.7%) | 1924 (70.0%) | 800 (71.7%) |
| **2** | 562 (24.3%) | 232 (23.2%) | 242 (28.1%) | 169 (27.3%) | 666 (24.2%) | 274 (24.6%) |
| **3 (Least Deprived)** | 118 (5.1%) | 43 (4.3%) | 46 (5.3%) | 25 (4.0%) | 158 (5.7%) | 41 (3.7%) |

Overall, we observed 148 distinct pairwise combinations of co-prescribed antipsychotics among the 25 antipsychotics identified from the records in this study. Of the most commonly occurring pairs of co-prescribed antipsychotics, we observed 1,109 instances of co-prescribed olanzapine and risperidone, 801 of aripiprazole and risperidone, and 788 of aripiprazole and olanzapine (Supplementary Figure 5).

Among patients who were ever prescribed antipsychotics during the study period, 3,313 (24.7%) had recorded measurements of QTc, 1,481 (11.1%) had measurements of prolactin, and 3,863 (28.8%) had measurements of BMI. In total, we identified 15,612 measurements of QTc, 2,971 measurements of prolactin level, and 11,234 measurements of BMI. The number of these measurements and whether or not these measurements represent adverse drug events are indicated in Table 2, stratified by whether or not these measurements were taken during or following APP within respective exposure periods.

**Table 2.** Clinical measurements within exposure period. Number of QTc, prolactin level, and BMI measurements within normal clinical ranges and how many were defined as adverse drug outcomes (i.e. prolonged QTc, hyperprolactinaemia, or overweight/obese) and whether or not these measurements occurred during or after a period of APP within the defined exposure period. Note that individuals may have both normal or abnormal clinical measurements during either exposure or non-exposure periods, as shown in Supplementary Figure 1.

| Clinical Measurement | Phenotype | Measured Outside Exposure Period | Measured Within Exposure Period |
| --- | --- | --- | --- |
| **QTc** | **Normal QTc** | 7525 (91.2%) | 6708 (91.1%) |
|  | **Prolonged QTc** | 724 (8.8%) | 655 (8.9%) |
| **Prolactin** | **Normal** | 787 (55.6%) | 568 (36.5%) |
|  | **Hyperprolactinaemia** | 629 (44.4%) | 987 (63.5%) |
| **BMI** | **Not Overweight or Obese** | 3094 (48%) | 1891 (39.5%) |
|  | **Overweight or Obese** | 3354 (52%) | 2895 (60.5%) |

The results of univariable and multivariable multilevel mixed-effects logistic regression are shown in Table 3. Controlling for all covariates, APP was associated with increased rates of hyperprolactinaemia (adjusted odds ratio 2.46; 95% C.I. 1.87-3.24) within the period of APP prescribing and up to 30 days after. APP was also associated with increased registration of BMI measurement in the overweight or obese range(adjusted odds ratio 1.75; 95% C.I. 1.33-2.31), within the period of APP prescribing and up to 6 months after. We did not find evidence that APP was associated with greater frequencies of registered observations of prolonged QTc (adjusted odds ratio 0.96; 95% C.I. 0.73-1.27), during or within 7 days after APP.

**Table 3.** Results of unadjusted and adjusted multilevel mixed-effects logistic regressions for prolonged QTc, hyperprolactinaemia, and BMI >25. Adjusted regressions are adjusted for number of measurements, age, gender, ethnicity, and IMD tertile. Coefficients are presented as odds ratios (ORs) with 95% confidence intervals. Statistically significant results (p<0.05) are indicated with an asterisk (*).

|  | Prolonged QTc |  | Hyperprolactinaemia |  | Overweight/Obese BMI |  |
| --- | --- | --- | --- | --- | --- | --- |
|  | Unadjusted OR | Adjusted OR | Unadjusted OR | Adjusted OR | Unadjusted OR | Adjusted OR |
| **Antipsychotic Polypharmacy** |  |  |  |  |  |  |
| **No** | 1 | 1 | 1 | 1 | 1 | 1 |
| **Yes** | 1.03<br>(0.78, 1.36) | 0.96<br>(0.73, 1.27) | 2.66*<br>(2.03, 3.48) | 2.46*<br>(1.87, 3.24) | 1.87*<br>(1.43, 2.45) | 1.75*<br>(1.33, 2.31) |
| **Number of Measurements** | 0.96<br>(0.92, 1.00) | 0.97<br>(0.93, 1.01) | 1.08*<br>(1.02, 1.15) | 1.03<br>(0.97, 1.09) | 0.97<br>(0.93, 1.01) | 0.94*<br>(0.90, 0.98) |
| **Age** | 1.04*<br>(1.04, 1.04) | 1.03*<br>(1.02, 1.05) | 0.98*<br>(0.97, 0.99) | 0.98*<br>(0.97, 0.99) | 1.01*<br>(1.00, 1.03) | 1.02*<br>(1.00, 1.03) |
| **Gender** |  |  |  |  |  |  |
| **Male** | 1 | 1 | 1 | 1 | 1 | 1 |
| **Female** | 0.45*<br>(0.24, 0.84) | 0.22*<br>(0.12, 0.40) | 1.97*<br>(1.44, 2.71) | 2.03*<br>(1.49, 2.77) | 0.54*<br>(0.36, 0.81) | 0.51*<br>(0.33, 0.78) |
| **Ethnicity** |  |  |  |  |  |  |
| **White British** | 1 | 1 | 1 | 1 | 1 | 1 |
| **White Other** | 0.81<br>(0.37, 1.76) | 0.37*<br>(0.18, 0.77) | 1.42<br>(0.93, 2.17) | 1.48<br>(0.98, 2.22) | 2.62*<br>(1.46, 4.69) | 1.10<br>(0.47, 2.58) |
| **Asian** | 1.03<br>(0.34, 3.11) | 0.10*<br>(0.02, 0.54) | 1.28<br>(0.66, 2.50) | 1.05<br>(0.55, 2.01) | 1.38<br>(0.59, 3.23) | 6.61*<br>(3.73, 11.69) |
| **Black** | 0.40*<br>(0.17, 0.91) | 0.28*<br>(0.13, 0.59) | 2.18*<br>(1.45, 3.28) | 1.71*<br>(1.15, 2.55) | 9.44*<br>(5.23, 17.03) | 3.19*<br>(1.10, 9.23) |
| **Mixed** | 0.47<br>(0.09, 2.41) | 0.04*<br>(0.00, 0.65) | 2.25*<br>(1.11, 4.58) | 1.62<br>(0.81, 3.24) | 3.56*<br>(1.23, 10.35) | 1.22<br>(0.48, 3.10) |
| **Other** | 0.57<br>(0.14, 2.37) | 0.27<br>(0.06, 1.11) | 1.01<br>(0.51, 2.00) | 0.93<br>(0.47, 1.81) | 1.68<br>(0.66, 4.24) | 2.24*<br>(1.26, 3.96) |
| **IMD Tertile** |  |  |  |  |  |  |
| **1 (Most Deprived)** | 1 | 1 | 1 | 1 | 1 | 1 |
| **2** | 1.3<br>(0.66, 2.57) | 0.69<br>(0.36, 1.34) | 0.94<br>(0.66, 1.33) | 0.99<br>(0.71, 1.39) | 0.54*<br>(0.33, 0.87) | 0.48*<br>(0.29, 0.80) |
| **3 (Least Deprived)** | 1.48<br>(0.39, 5.64) | 0.11*<br>(0.01, 0.99) | 2.22*<br>(1.04, 4.73) | 2.5*<br>(1.21, 5.2) | 0.13*<br>(0.05, 0.34) | 0.14*<br>(0.05, 0.38) |

Female patients, adjusting for all covariates, had an increased odds of hyperprolactinaemia (adjusted odds ratio 2.03; 95% C.I. 1.49-2.77) compared to men, and fewer instances of both prolonged QTc (adjusted odds ratio 0.22; 95% C.I. 0.12-0.40) and overweight/obesity (adjusted odds ratio 0.51; 95% C.I. 0.33-0.78). As compared to White British patients and adjusting for all covariates, Black patients were more likely to experience both hyperprolactinaemia (adjusted odds ratio 1.71; 95% C.I. 1.45-3.28) and a BMI measurement consistent with being overweight or obese (adjusted odds ratio 3.19; 95% C.I. 1.10-9.23).

## Discussion

### Key Findings

Using routinely collected EHR data, we observed that approximately a quarter (24.7%) of all patients prescribed antipsychotics from 2008-18 showed at least one period of APP at C&I NHS FT. We observed aripiprazole, olanzapine, and risperidone as among the most commonly co-prescribed antipsychotics while benperidol, lurasidone, pimozide, promazine, and ziprasidone were the least commonly co-prescribed antipsychotics. We found increased risk of hyperprolactinaemia during or up to 30 days following APP, and BMI of 25 or greater during or up to 180 days following APP. We found no association between APP and subsequent measures of prolonged QTc.

### Limitations and Strengths

While the extensive EHR data at C&I NHS FT have allowed us to explore the temporal relationship between APP prescribing and three commonly associated ADRs related to antipsychotic medication, there are also limitations to this approach. Firstly, all studies relying upon routinely collected data, such as EHR data, are limited to data which are collected and recorded based on routine care. This necessarily means that many measurements which would normally be included in a conventional study design may not be present in the EHR, either because they were not taken or not recorded. Moreover, because EHR data does not capture information on whether patients fill their prescriptions or whether patients actually take their medications, it may be possible, despite mentions of antipsychotics in EHR records, that patients may not actually be taking antipsychotics as prescribed. For example, in December 2020, 97,149,986 items were prescribed in England (NHS Business Services Authority, n.d.), whereas only 89,592,218 items (92%) were dispensed (NHS Business Services Authority, n.d.). Analyses of other sources of routinely collected data, such as claims or pharmacy dispensing data, can potentially provide further validation yet these methods can also be subject to inaccuracy, particularly without clinical confirmation of patient adherence to prescribed antipsychotic(s). In addition, while it is possible to examine specific dosages of prescribed antipsychotics, this was not included in our study at present due to the added complexity this would add to our analysis model. Future approaches should consider the impact of dosage with respect to both APP and monotherapy, as high-dose monotherapy may present similar risks as APP. In addition, future research, should examine the effects of medication delivery, specifically testing if oral only polypharmacy vs mixed oral/depot (long-acting injections) has differential impact on ADRs. In our current analysis any combination of more than one antipsychotic drug given over a period of 30 days was considered polypharmacy regardless of the mode of delivery. Moreover, this study is limited by the quality of routinely collected EHR and by the fact that this data was not collected for research purposes. As such, variations in free-text clinical documentation and notes may result in per-individual differential recording of antipsychotic prescribing, complicating attempts to identify instances through NLP. As the data are routinely, but potentially not regularly, collected, there may be gaps in recorded antipsychotic prescribing for which this study has not accounted. Prescriptions from sources external to C&I NHS FT such as GPs and/or out of borough consultations will also be missing.

Similarly, gaps in surveillance of ADRs or differential examination among patients could also affect our results, and this is a notable limitation when analysing routinely collected data from EHR. To partially account for potential bias arising from some patients receiving frequent treatment potentially undergoing more tests for ADRs than other patient groups, we included the total number of measures as a covariate in our analysis. In addition, the time to ADR onset following APP initiation may be very short and potentially missed if measurements do not occur within a specific observation window; for instance, lack of timely electrocardiogram (ECG) measurements or their recording in the EHR may mean that some instances of QTc prolongation may have been overlooked in our analysis.

Nevertheless, our findings are limited insofar that we cannot eliminate the non-random error which may be introduced from lack of randomization. It’s also unclear to what extent our observed results might be affected by carry over effects, where earlier treatments could have impacted on subsequent overweight or hyperprolactinaemia.

Notwithstanding, the use of routinely collected data offers the opportunity to conduct large-scale studies with real-world data without the need for extensive measurement and testing. This approach can provide initial guidance for further research and input to clinical practice, which is essential given ours and others observations that APP is common in the clinical setting (Kadra *et al*., 2015).

#### Comparison to Prior Findings

Our findings are broadly consistent with previous literature, although we note that prior attempts to systematically synthesize findings across heterogeneous studies investigating the safety of APP have sometimes been inconclusive (Gallego *et al*., 2012; Fleischhacker and Uchida, 2014; Takeuchi *et al*., 2015; Kadra *et al*., 2018b, 2018a; Patel *et al*., 2018; Taipale *et al*., 2018; Buhagiar *et al*., 2020). Further research utilising comparable datasets would strengthen the evidence to inform the use of APP in the clinical setting. Our use of real-world EHR data shows promise in advancing the knowledge in this area.

#### Future Directions

This study provides evidence that routinely collected EHR data can be effectively used to conduct pharmacoepidemiological studies to inform clinical practice, even in the absence of pharmacy dispensing data. The quality and volume of EHR data continues to increase, particularly as these data contribute to care quality improvement initiatives. As such, EHR research offers many opportunities to address pressing research questions which have proven difficult to address through traditional research means, such as RCTs. Future work may look at the impact of dosage, the effects of increasing number of concurrently prescribed antipsychotics during APP (e.g. the extend and effects of three or more antipsychotics being co-prescribed at one time), and/or the distinction between oral and depot prescriptions of antipsychotics. In addition, further linkages with complementary datasets, such as primary care records and civil mortality registers, and work to develop and validate additional NLP algorithms may permit a fuller examination of patient side effect outcomes, such as hemoglobin A1c, lipoproteins, diabetes and other important clinical outcomes not examined in this study. Furthermore, to address the non-random error which may be introduced by the lack of randomization in analysing routinely collected data, other study designs such as stratified Cox regression or self-controlled case series may offer methodologically advantageous approaches for further analyses.

The study also contributes to the growing body of evidence that APP should be carefully managed in the clinical setting with a particular focus on the monitoring of prolactin and weight gain, and other physical health indicators, especially given the common occurrence of APP in the clinical setting.

## Data Availability

Data used in this article can only be access by approved researchers, using the C&I CRIS research database. If you would like further information about using CRIS at C&I for a project, please contact the C&I Research Database Manager by emailing.
If you would like to submit an application for permission to use CRIS for a research project, please download the project application form here.

https://www.candi.nhs.uk/health-professionals/research/ci-research-database

