## Supplementary Material for "Antipsychotic Polypharmacy and Adverse Drug Reactions Among Adults in a London Mental Health Service, 2008-2018"

**Supplementary Table 1.** Antipsychotics identified in this study

| Antipsychotic | Observations | Unique Patients | Median Duration (Days) |
|---|---|---|---|
| amisulpride | 1,674 | 1,219 | 301.0 |
| aripiprazole | 4,017 | 2,794 | 178.0 |
| benperidol | <10 | <10 | 56.0 |
| chlorpromazine | 690 | 551 | 243.0 |
| clozapine | 2,834 | 1,127 | 173.0 |
| flupentixol | 1,173 | 940 | 547.0 |
| fluphenazine | 158 | 127 | 1,010.0 |
| haloperidol | 242 | 207 | 445.0 |
| levomepromazine | 47 | 39 | 42.0 |
| lurasidone | 15 | 11 | 31.0 |
| olanzapine | 8,330 | 5,423 | 185.0 |
| paliperidone | 326 | 264 | 185.0 |
| periciazine | <10 | <10 | 94.5 |
| perpenazine | <10 | <10 | 52.0 |
| pimozide | <10 | <10 | 93.0 |
| pipotiazine | 226 | 190 | 520.0 |
| prochlorperazine | 97 | 97 | 111.0 |
| promazine | 10 | 10 | 69.0 |
| quetiapine | 5,088 | 3,741 | 161.0 |
| risperidone | 8,084 | 5,132 | 180.0 |
| sulpride | 591 | 501 | 307.0 |
| thioridazine | 18 | 18 | 304.0 |
| trifluoperazine | 181 | 146 | 269.0 |
| ziprasidone | <10 | <10 | 70.0 |
| zuclopenthixol | 1,588 | 1,165 | 359.5 |

All EHR free-text was converted to lower case and the following expressions were used to identify our ADR measures.

**Supplementary Table 2.** Regular expressions used to identify and extract clinical measurements from clinical free-text.

<table>
<tr><th>Clinical Measure</th><th>Regular Expression</th><th>Examples of matches</th><th>PPV</th><th>NPV</th></tr>
<tr><td>QTc</td><td>qtc ?[a-z-=:–]{0,3} ?([0-9/]+)[ms.]*</td><td>qtc – 416ms, "qtc 433", "qtc is 432", "qtc:456", "qtc-546", "qtc=546", "qtc=546ms.", "qtc=546ms"</td><td>100%</td><td>100%</td></tr>
<tr><td>Prolactin</td><td>[ ]?prolactin[* ]*([0-9]+) *[-0-9\\[\\]\\(\\) ]* *[muil\\/]+ *[-0-9\\[\\]\\(\\) ]*</td><td>Prolactin 97 mu/L 86 - 324, " Prolactin * 603 mu/L [86 - 324] ", "Prolactin 264 102-496 mIU/L", "Prolactin * 1317 mu/L 102 - 496", "Prolactin 128 mu/L 86 - 324", "Prolactin * 603 mu/L 86 - 324"</td><td>100%</td><td>100%</td></tr>
<tr><td>BMI</td><td>[ ]?(bmi|body mass index) ?([=:–-]|around|which is|is)? *([0-9]+\\.?[:digit:]{0,2})</td><td>- BMI: 28.68 -,"BMI 32", "BMI= 65", " BMI 33.9 ", "BMI 26", " BMI which is 17.5 an", "BMI = 27kg/", "(BMI:22kg/m2)", "BMI – 22( ", " BMI around 21 ", "BMI = 23.4kg/m2.", "13 Body mass index 25.2 ", "Body mass Index 25.2 "</td><td>100%</td><td>100%</td></tr>
</table>

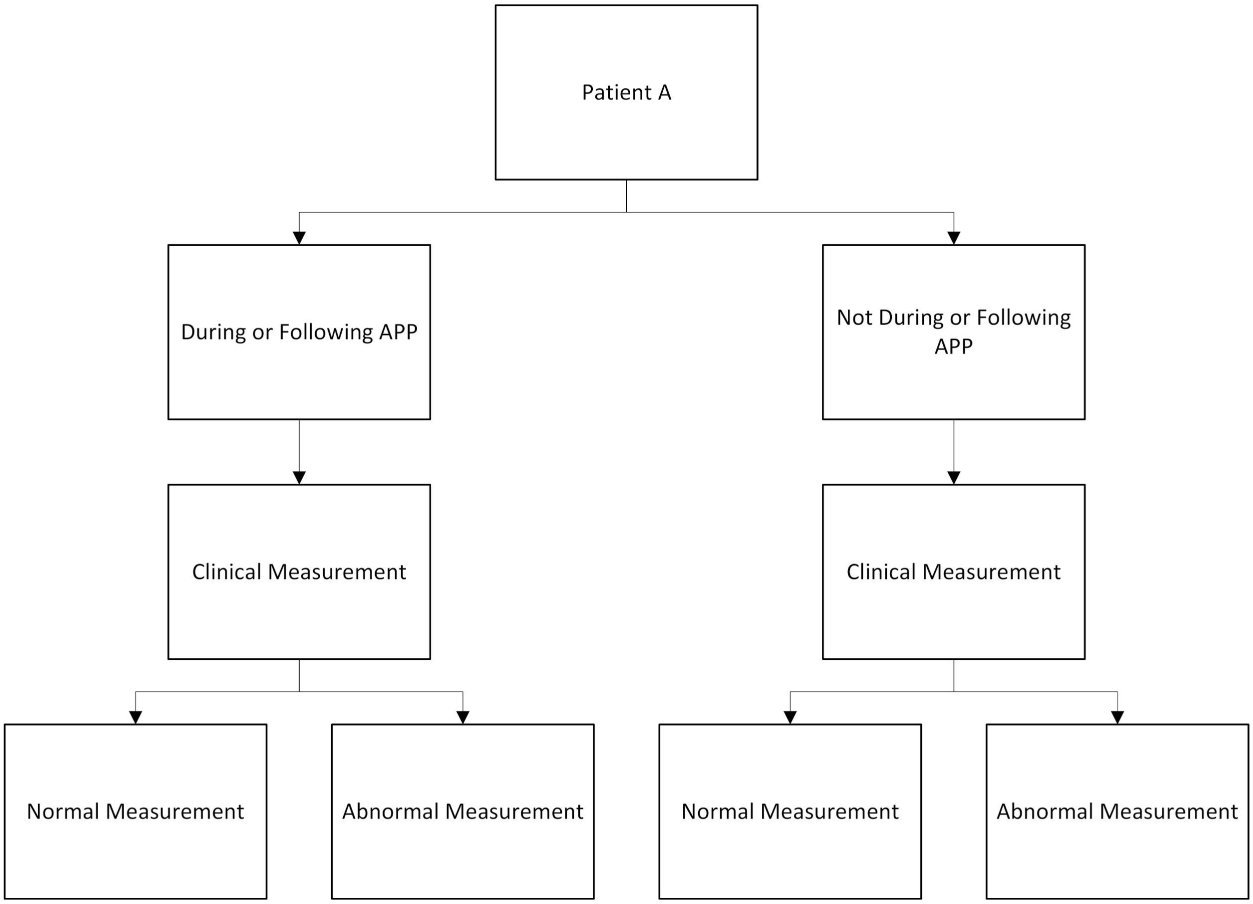


**Supplementary Figure 1.** Schematic of clinical measurements identified per individual patient. Multilevel mixed-effects modeling with the patient as the random effects, means that we can include all four bottom row states for each individual (should they exist at different timepoints), accounting for within-patient clustering of measurements, in our analysis of the effects of antipsychotic polypharmacy on adverse drug reactions.

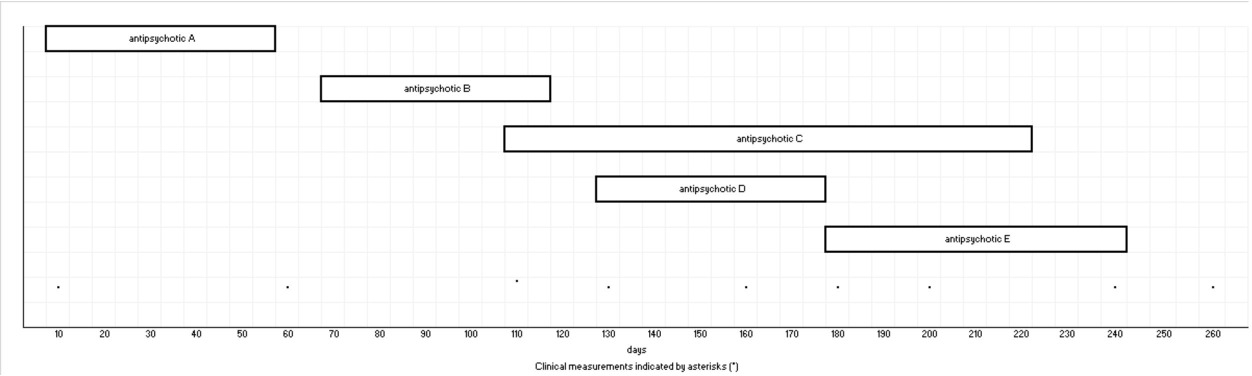


**Supplementary Figure 2**. Schematic of polypharmacy periods and classification of clinical measurements for a single hypothetical patient. Clinical measurements are denoted by asterisks (*). In this example, clinical measurements on day 10, day 60, day 110 would not be considered during APP exposure periods. The clinical measurement on day 10 is during a period of antipsychotic monotherapy. The clinical measurement on day 60 is not during any period of any antipsychotic prescribing. The measurement on day 110 does not occur during a period of APP because antipsychotic B and antipsychotic C are coprescribed for less than 30 days. Clinical measurements on day 130, 160, 180, 200, and 240 would be considered during APP exposure periods. The clinical measurement on day 260 would be considered outside an APP exposure period if it were a measurement of QTc but would be considered within an APP exposure period if it were a measurement of either prolactin or BMI. All clinical measurements for this patient would be included in this study.

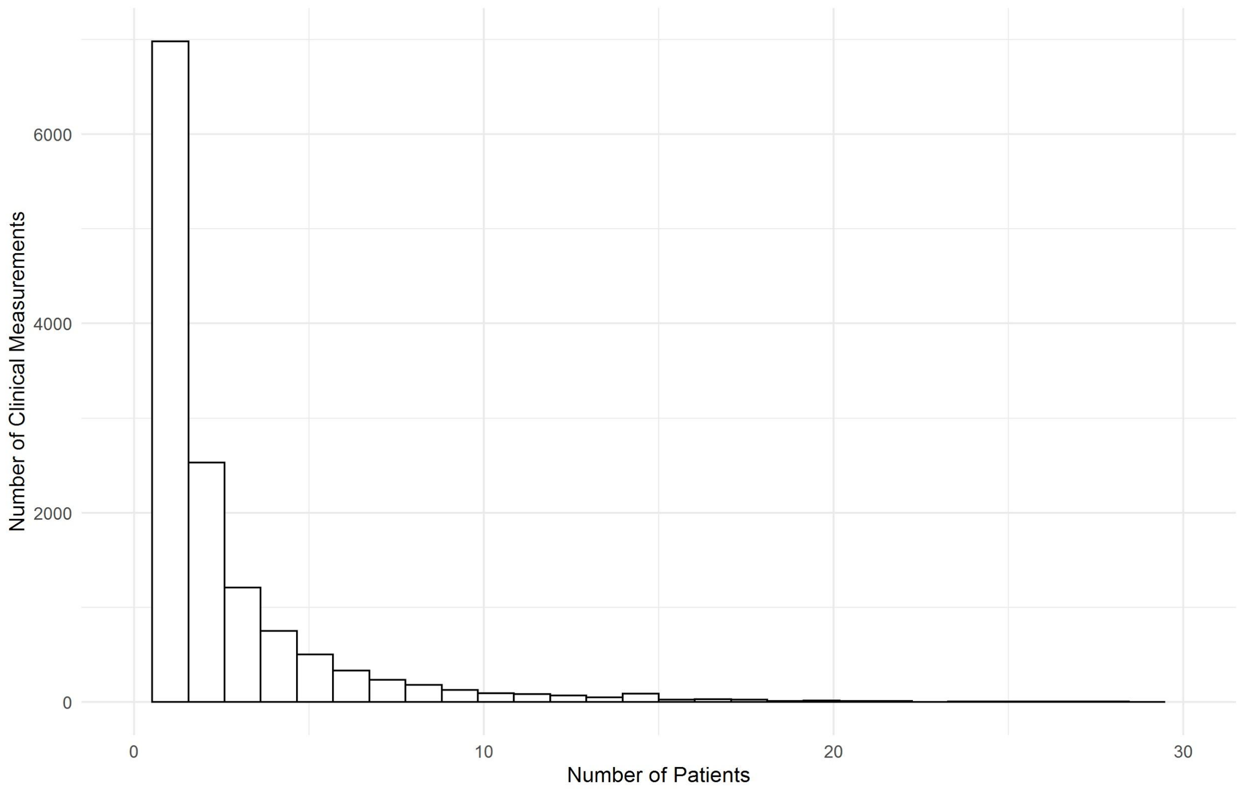


**Supplementary Figure 3.** Distribution of clinical test of adverse drug reaction per patient

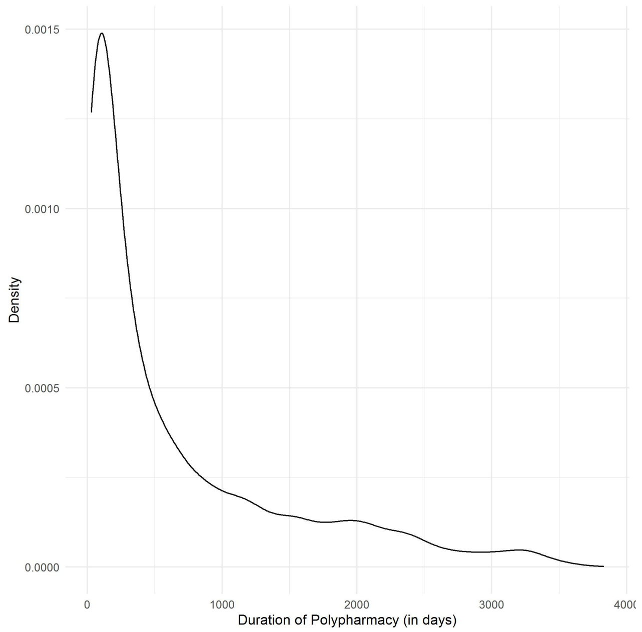


**Supplementary Figure 4.** Density plot of duration of periods of antipsychotic polypharmacy (in days)

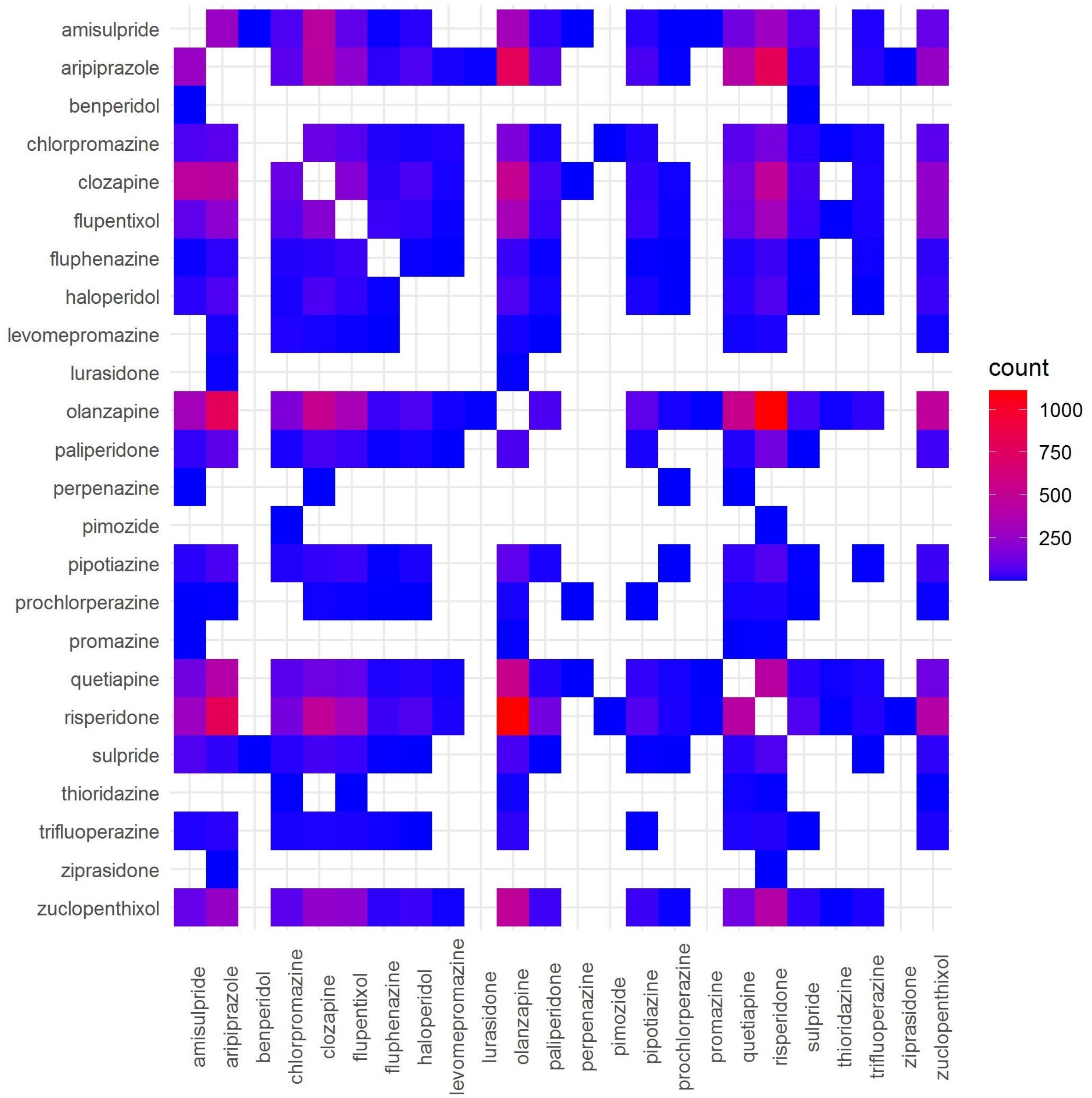


**Supplementary Figure 5**. Heatmap indicating pairwise frequencies of co-prescribed antipsychotics. Empty spaces indicate no observed pairwise co-prescription.
